# Can Routine Clinical Variables Substitute for Urine Albumin Testing in Moderate Chronic Kidney Disease? A Machine-Learning Analysis of NHANES 2015–2018

**DOI:** 10.64898/2026.04.03.26349862

**Authors:** Jaswant Narendra Saxena, Devi Vara Prasad Potturu, Ananya Nagraj

**Affiliations:** CGI Technologies and Solutions Inc., United States; Indian School of Business, Hyderabad, India; Department of Management Information Systems, Stevens Institute of Technology, Hoboken, NJ, USA

**Keywords:** chronic kidney disease, albuminuria, urine albumin-to-creatinine ratio, machine learning, logistic regression, NHANES, screening

## Abstract

**Background:** Albuminuria is central to staging and risk prediction in chronic kidney disease (CKD), yet urine albumin-to-creatinine ratio (UACR) testing is performed far less often than serum creatinine testing. Whether routinely available clinical variables can identify patients likely to have albuminuria, and so substitute for or prioritize testing, is unclear.

**Methods:** We analyzed adults in NHANES 2015–2018 with an eGFR of 30–59 mL/min/1.73m^2^ (CKD-EPI 2021). The outcome was UACR *≥*30 mg/g. Seventeen features derived from demographics, serum creatinine, eGFR, blood pressure, diabetes status and self-reported kidney disease were used; UACR was excluded. Logistic regression, random forest and gradient boosting were evaluated with 10-times repeated stratified 5-fold cross-validation. We report AUC with bootstrap 95% CIs, DeLong tests, calibration, survey-weighted AUC, diabetes subgroups and a rule-out analysis.

**Results:** Of 687 participants, 239 (34.8%) had UACR *≥*30 mg/g. Discrimination was modest and similar across models: logistic regression AUC 0.723 (95% CI 0.684–0.762), random forest 0.721 (0.680–0.761; DeLong *p* = 0.84 vs logistic regression) and gradient boosting 0.719 (0.679–0.759; *p* = 0.76). Logistic regression and random forest were reasonably calibrated (slopes 0.87 and 0.98); gradient boosting was overfitted (slope 0.64). Discrimination was lower in participants without diabetes (logistic regression AUC 0.636 vs 0.722 with diabetes). Using logistic regression to rule out testing at 90% sensitivity would have spared 22.7% of participants from testing but missed 23 of 239 cases; at 95% sensitivity, only 16.3% would have been spared.

**Conclusions:** Routine clinical variables identify albuminuria only modestly, and flexible machine-learning models offer no advantage over logistic regression. No model allowed a meaningful share of patients to forgo UACR testing without missing cases, supporting universal UACR testing in moderate CKD.

## 1. Introduction

Chronic kidney disease (CKD) affects an estimated 850 million people worldwide [1, 2], and about 14% of US adults, roughly 37 million people [3]. Guidelines stage CKD by both estimated glomerular filtration rate (eGFR) and albuminuria, because albuminuria independently predicts kidney failure, cardiovascular events and death [4]. Albuminuria is also an input to the Kidney Failure Risk Equation (KFRE) [5], which has been validated in more than 700,000 participants across 31 cohorts [6].

Despite this, albuminuria is under-tested. In a US national laboratory database of 28.3 million adults with diabetes or hypertension, only 21.0% received a urine albumin-to-creatinine ratio (UACR) test in a year, compared with 89.6% who had eGFR testing [7]. In an international consortium, albuminuria screening rates were 35.1% in diabetes and 4.1% in hypertension, and most albuminuria in people with hypertension was estimated to be undetected [8].

Because serum creatinine, blood pressure and diabetes status are almost always available, it is tempting to ask whether these variables, perhaps combined by machine learning, could identify who is likely to have albuminuria. Such a model could, in principle, either replace testing or prioritize it. Machine-learning studies have predicted proteinuria and diabetic kidney disease from electronic health records [9, 10], but systematic evidence shows that flexible machine-learning methods often perform no better than logistic regression for clinical prediction [11].

We therefore asked three questions in a nationally representative US sample of adults with moderately reduced eGFR: (1) how well can routine clinical variables, without UACR, identify albuminuria; (2) do machine-learning models outperform logistic regression; and (3) could any model safely reduce the need for UACR testing?

## 2. Methods

### 2.1 Data source

The National Health and Nutrition Examination Survey (NHANES) is a cross-sectional survey of the civilian, non-institutionalized US population. We pooled the 2015–2016 and 2017–2018 cycles and merged six public-use modules by respondent sequence number: Demographics, Standard Biochemistry Profile, Albumin and Creatinine (urine), Blood Pressure, Diabetes questionnaire and Kidney Conditions questionnaire. NHANES protocols were approved by the NCHS Research Ethics Review Board; this secondary analysis used de-identified public data. Reporting follows the TRIPOD+AI statement where applicable [12].

### 2.2 Study population

We included adults aged *≥*18 years with serum creatinine available and an eGFR of 30–59 mL/min/1.73m^2^, calculated with the race-free CKD-EPI 2021 creatinine equation [13]. We excluded participants without a valid UACR, those reporting dialysis in the previous 12 months and pregnant participants. Of 19,225 participants in the merged files, 11,848 were adults, 10,539 had serum creatinine, 729 had an eGFR of 30–59, 689 had UACR available and 687 remained after excluding dialysis (no participant was excluded for pregnancy). Eligibility was based on a single eGFR measurement, so the cohort approximates, but is not identical to, clinically confirmed CKD G3, which requires persistence for more than three months [4].

### 2.3 Outcome

The outcome was albuminuria, defined as UACR *≥*30 mg/g (KDIGO category A2 or A3) [4], using the NHANES variable URDACT from a single spot urine sample.

### 2.4 Predictors

Seventeen predictors were pre-specified: age; sex; diabetes (DIQ010 = 1); pre-diabetes (DIQ010 = 3); serum creatinine and its square; eGFR and its square; an indicator for eGFR *<*45; mean systolic and diastolic blood pressure (up to three readings; diastolic readings recorded as 0 treated as missing); pulse pressure; systolic pressure *>*140 mmHg; self-reported kidney disease (KIQ022 = 1); and the interactions age *×* diabetes, eGFR *×* diabetes and age *×* eGFR. UACR was not used as a predictor. Missingness was low (blood pressure variables 2.9%, self-reported kidney disease 0.9%, diabetes 0.1%) and was handled by median imputation fitted within each training fold.

### 2.5 Models

We compared three models: logistic regression with L2 regularization (*C* = 1.0) after standardization; random forest (500 trees, maximum depth 6, minimum 10 samples per leaf); and gradient boosting (300 trees, maximum depth 3, learning rate 0.05, subsampling 0.8, minimum 10 samples per leaf). Hyperparameters were fixed in advance rather than tuned, to limit optimism in a small sample. All models were implemented in scikit-learn [14].

### 2.6 Validation and statistical analysis

Performance was estimated by stratified 5-fold cross-validation repeated 10 times; imputation and scaling were fitted only on training folds. For each participant, out-of-fold predicted probabilities were averaged across the 10 repetitions. Discrimination was summarized by the area under the ROC curve (AUC) with 95% CIs from 2,000 bootstrap resamples, and models were compared with logistic regression using DeLong’s test [15, 16]. Calibration was assessed with calibration plots by decile, calibration intercept and slope, and the Brier score [17]. Average precision was also reported. As a sensitivity analysis, AUC was recomputed using the NHANES 4-year mobile examination center weights to approximate population-level performance.

To translate discrimination into clinical terms, we performed a rule-out analysis: for each model, we chose the probability threshold that achieved 90% or 95% sensitivity and calculated the share of participants below the threshold (who could, hypothetically, skip UACR testing), the number of albuminuria cases they included and the negative predictive value. Discrimination was also estimated separately for participants with and without diabetes. Predictor importance was described with standardized logistic regression coefficients and permutation importance for gradient boosting; these were computed on models fitted to the full data and are descriptive.

Analyses used Python with scikit-learn 1.6.1, pandas 2.2.3 and NumPy 2.1.3, with a fixed random seed (42). The full code, which downloads the data directly from the CDC, is openly available on Zenodo [18] (https://doi.org/10.5281/zenodo.23267842).

## 3. Results

### 3.1 Participant characteristics

Of 687 participants, 239 (34.8%) had UACR *≥*30 mg/g (Table 1). Mean age was 71.0 years, 50.9% were female and 35.1% had diagnosed diabetes. Compared with participants without albuminuria, those with albuminuria more often had diabetes (54.2% vs 25.0%), had higher systolic blood pressure (144.2 vs 133.0 mmHg) and lower eGFR (47.0 vs 50.6 mL/min/1.73m^2^), and more often reported having been told they had kidney disease (30.8% vs 17.7%). Age was similar (71.1 vs 70.9 years).

**Table 1:** Participant characteristics by albuminuria status, NHANES 2015–2018 adults with eGFR 30–59 mL/min/1.73m^2^.

| Characteristic | All ($n = 687$) | UACR $\geq 30$ ($n = 239$) | UACR $< 30$ ($n = 448$) |
| --- | --- | --- | --- |
| Age, years, mean (SD) | 71.0 (10.5) | 71.1 (10.8) | 70.9 (10.4) |
| Female, $n$ (%) | 350 (50.9) | 110 (46.0) | 240 (53.6) |
| Diabetes, $n$ (%) | 241 (35.1) | 129 (54.2) | 112 (25.0) |
| Pre-diabetes, $n$ (%) | 18 (2.6) | 7 (2.9) | 11 (2.5) |
| eGFR, mL/min/1.73m<sup>2</sup>, mean (SD) | 49.3 (8.0) | 47.0 (8.6) | 50.6 (7.5) |
| Serum creatinine, mg/dL, mean (SD) | 1.4 (0.3) | 1.4 (0.3) | 1.3 (0.3) |
| Systolic BP, mmHg, mean (SD) | 136.9 (23.2) | 144.2 (24.4) | 133.0 (21.6) |
| Systolic BP $> 140$ mmHg, $n$ (%) | 262 (39.3) | 121 (52.2) | 141 (32.4) |
| Told of kidney disease, $n$ (%) | 151 (22.2) | 72 (30.8) | 79 (17.7) |
| UACR, mg/g, median (IQR) | 15.2 (7.3–58.8) | 110.3 (56.8–397.5) | 9.0 (5.5–14.8) |

Percentages are among participants with non-missing data. Unweighted estimates. BP, blood pressure.

### 3.2 Discrimination and calibration

All three models showed modest, nearly identical discrimination (Table 2; Figure 1). Logistic regression achieved an AUC of 0.723 (95% CI 0.684–0.762); random forest and gradient boosting did not differ from it (differences *−*0.002 and *−*0.004; DeLong *p* = 0.84 and 0.76). Survey-weighted AUCs were similar (0.709–0.722). Logistic regression and random forest were reasonably calibrated (calibration slopes 0.87 and 0.98), whereas gradient boosting produced predictions that were too extreme (slope 0.64), indicating overfitting (Figure 2).

**Table 2:** Cross-validated performance for identifying UACR *≥*30 mg/g (687 participants, 239 cases).

| Model | AUC (95% CI) | ΔAUC vs LR | DeLong $p$ | AP | Brier | Cal. slope | Weighted AUC |
| --- | --- | --- | --- | --- | --- | --- | --- |
| Logistic regression | 0.723 (0.684–0.762) | — | — | 0.591 | 0.193 | 0.87 | 0.722 |
| Random forest | 0.721 (0.680–0.761) | −0.002 | 0.84 | 0.594 | 0.192 | 0.98 | 0.709 |
| Gradient boosting | 0.719 (0.679–0.759) | −0.004 | 0.76 | 0.600 | 0.199 | 0.64 | 0.711 |

5-fold cross-validation repeated 10 times; CIs from 2,000 bootstrap resamples. LR, logistic regression; AP, average precision; Cal. slope, calibration slope (ideal 1.0); Weighted AUC, AUC using NHANES examination weights. Calibration intercepts were 0.00, 0.01 and 0.06.

**Figure 1:**
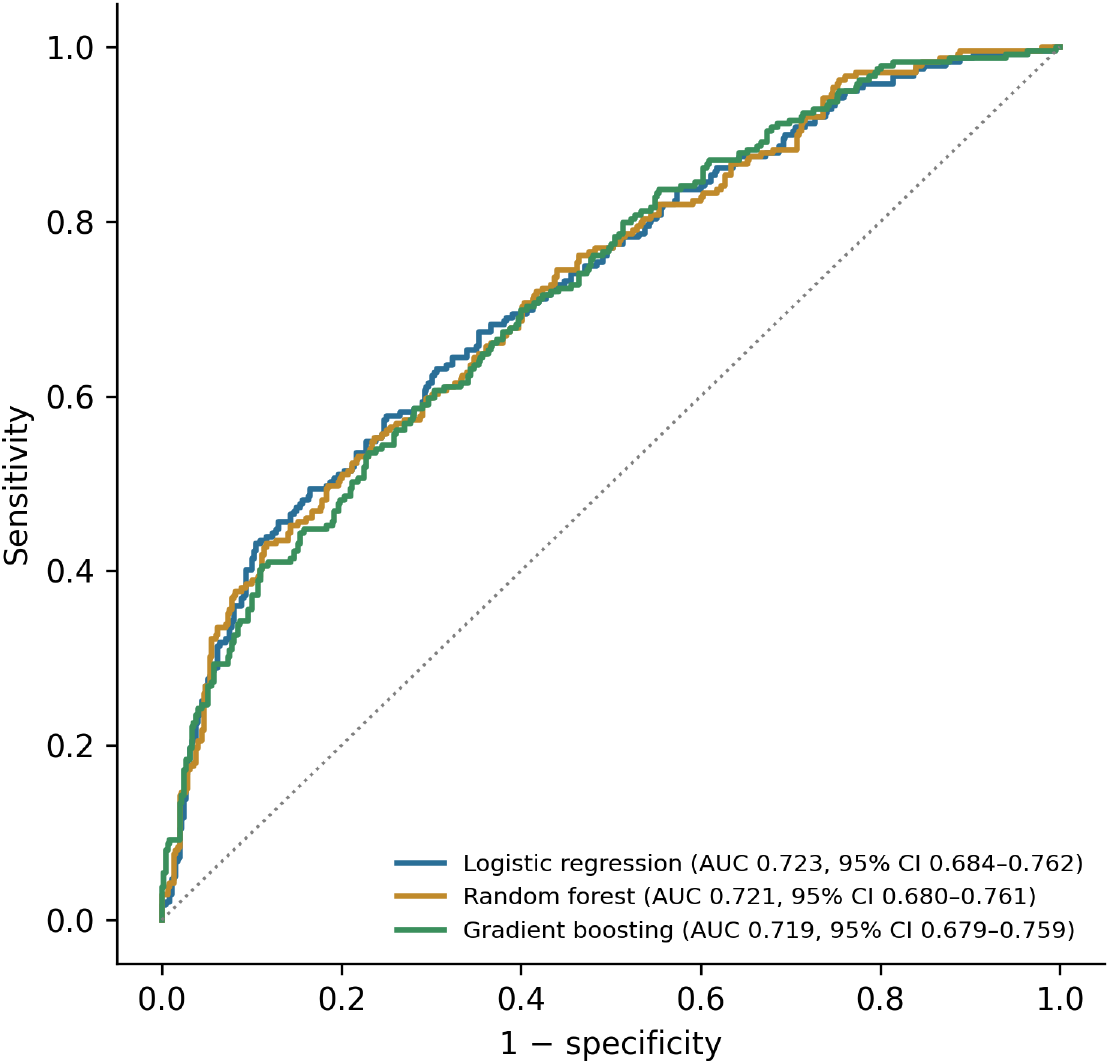
Cross-validated ROC curves for identifying UACR *≥*30 mg/g from routine clinical variables.

**Figure 2:**
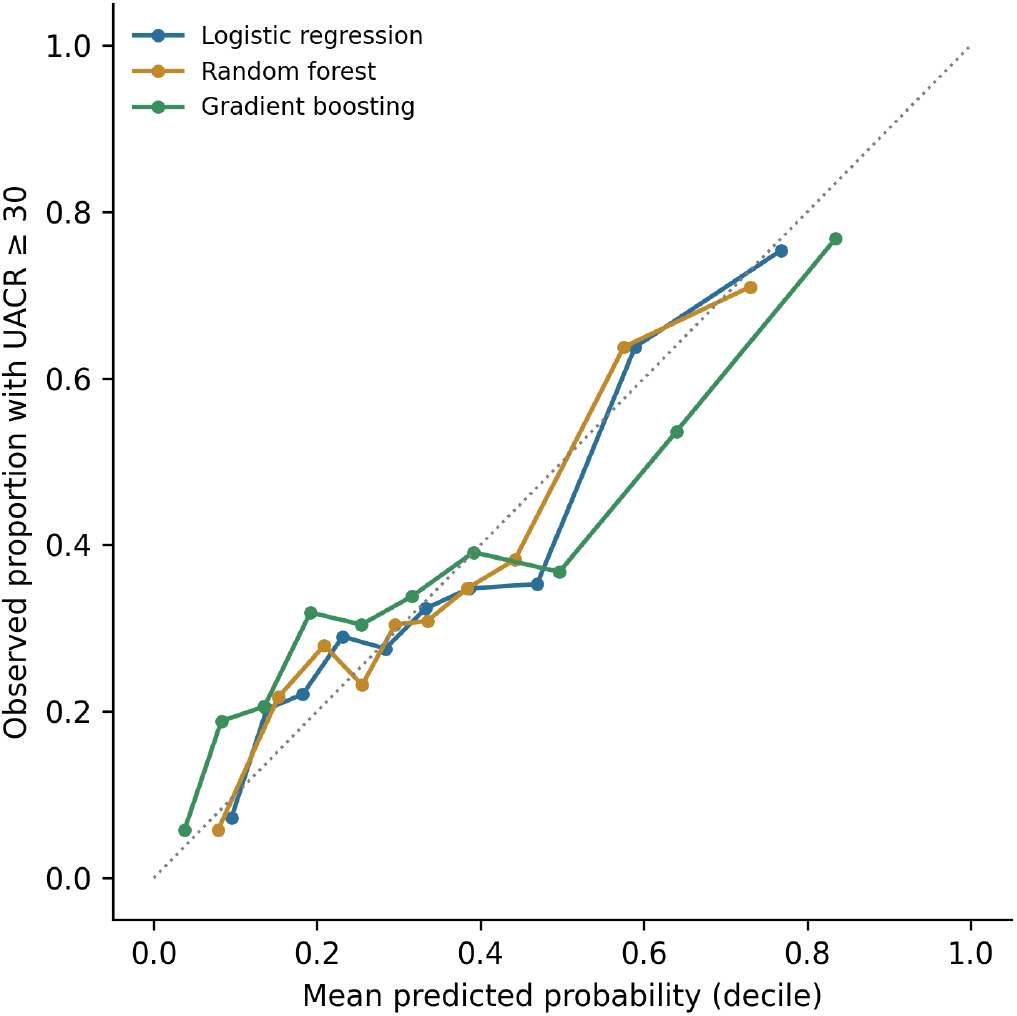
Calibration by decile of predicted probability. The dotted line indicates perfect calibration.

### 3.3 Rule-out analysis

No model could identify a large group at low enough risk to forgo testing (Table 3). With logistic regression at a threshold giving 90% sensitivity, 156 participants (22.7%) fell below the threshold, but 23 of them had albuminuria (negative predictive value 0.85), meaning that roughly 1 in 7 people spared testing would have had undetected albuminuria. At 95% sensitivity, only 112 participants (16.3%) fell below the threshold, of whom 11 had albuminuria (NPV 0.90). Results were similar for the other models.

**Table 3:** Rule-out analysis: share of participants who could hypothetically skip UACR testing at fixed sensitivity.

| Model | Target sensitivity | Below threshold, % | Cases missed (of 239) | NPV |
| --- | --- | --- | --- | --- |
| Logistic regression | 90% | 22.7 | 23 | 0.85 |
|  | 95% | 16.3 | 11 | 0.90 |
| Random forest | 90% | 22.3 | 23 | 0.85 |
|  | 95% | 18.0 | 11 | 0.91 |
| Gradient boosting | 90% | 24.6 | 23 | 0.86 |
|  | 95% | 16.4 | 11 | 0.90 |

Achieved sensitivities were 90.4% and 95.4%. NPV, negative predictive value among participants below the threshold.

### 3.4 Subgroups and predictors

Discrimination was higher among participants with diabetes (*n* = 241; albuminuria 53.5%; logistic regression AUC 0.722, 95% CI 0.655–0.786) than among those without diabetes (*n* = 445; albuminuria 24.5%; AUC 0.636, 0.581–0.695), with the same pattern for random forest (0.718 vs 0.616) and gradient boosting (0.706 vs 0.633). Blood pressure variables and diabetes-related terms were the strongest predictors: systolic blood pressure, the age *×* eGFR interaction, pulse pressure and diastolic pressure had the largest permutation importance in gradient boosting (Figure 3), and the age *×* diabetes and diabetes terms had the largest standardized coefficients in logistic regression.

**Figure 3:**
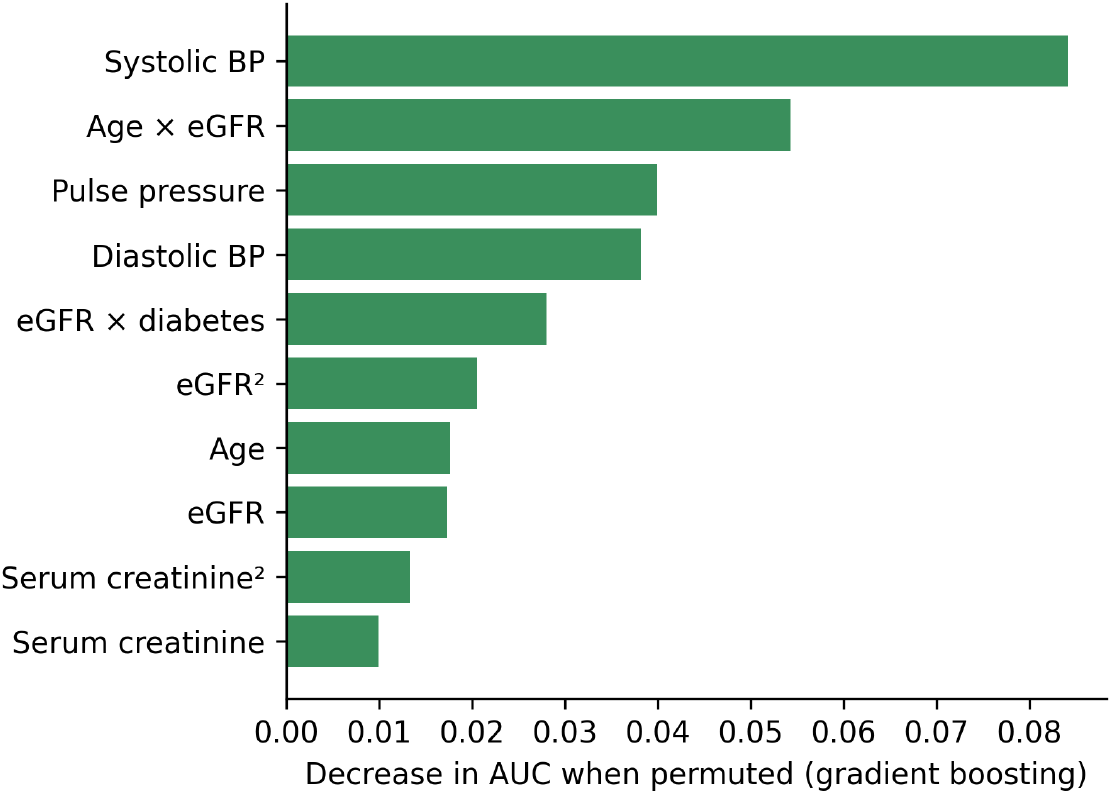
Ten most important predictors in gradient boosting (permutation importance, decrease in AUC). Computed on the full data; descriptive only.

## 4. Discussion

### 4.1 Principal findings

In a nationally representative sample of US adults with moderately reduced eGFR, routine clinical variables identified albuminuria with only modest discrimination (AUC about 0.72). Random forest and gradient boosting did not improve on logistic regression, and gradient boosting was poorly calibrated. Most importantly, no model could safely reduce testing: at 90% sensitivity, about one in five patients would have been spared testing, but one in seven of those would have had undetected albuminuria. Performance was worse in people without diabetes, the group in which albuminuria testing is already least common [8].

### 4.2 Interpretation

The limited discrimination is consistent with albuminuria and eGFR reflecting different aspects of kidney damage: albuminuria is associated with eGFR but adds independent information [4, 19]. Blood pressure and diabetes were the main predictors, in line with their known association with albuminuria [8], but these associations are too weak at the individual level to replace measurement.

The lack of benefit from machine learning agrees with a systematic review that found no average performance advantage for machine learning over logistic regression in clinical prediction [11]. With 239 events and largely linear relationships, flexible models have little structure to exploit and more room to overfit, as the calibration slope of 0.64 for gradient boosting shows. In settings like this, a well-specified logistic regression is a better default.

### 4.3 Implications

For health systems and value-based care programs that manage CKD populations, these results argue against using predictive models to decide who needs UACR testing. A model could still be useful for prioritizing outreach in a population that has not been tested, for example ordering who is contacted first, but every patient with eGFR below 60 should ultimately receive a UACR measurement, in line with guidelines [4]. Closing the testing gap [7] is likely to deliver more value than better prediction from the variables already collected.

### 4.4 Strengths and limitations

Strengths include the use of public, nationally representative data; a fully reproducible pipeline that downloads data directly from the CDC; fold-contained preprocessing; repeated cross-validation with confidence intervals, formal model comparison and calibration; and a rule-out analysis that expresses performance in clinically meaningful terms.

There are several limitations. First, both eGFR and UACR were measured once, so participants with transient changes may be misclassified; CKD and albuminuria categories formally require persistence for three months [4]. Second, the sample was modest (687 participants, 239 events), which limits precision, particularly in subgroups. Third, validation was internal; external validation in clinical or claims data is needed. Fourth, we did not include medications such as renin–angiotensin system inhibitors, body mass index, race and ethnicity, or other laboratory values, which could improve discrimination somewhat; however, the gap to a performance level that would allow safe rule-out is large. Fifth, models were trained without survey weights, and survey weights were applied only in a sensitivity analysis of discrimination. Finally, predictor importance was estimated in-sample and should be interpreted descriptively.

## 5. Conclusion

Among US adults with eGFR 30–59 mL/min/1.73m^2^, routine clinical variables identified albuminuria only modestly, and machine-learning models offered no improvement over logistic regression. No model allowed a meaningful share of patients to skip UACR testing without missing cases. Rather than predicting albuminuria, CKD care programs should focus on measuring it.

## Data Availability

The NHANES 2015 till 2018 data used in the primary analysis are publicly available at
https://wwwn.cdc.gov/nchs/nhanes. All analysis code, feature engineering pipelines,
and evaluation protocols will be made publicly available upon journal acceptance.
The NHANES-calibrated synthetic longitudinal cohort was generated using parameters
derived from the public NHANES data and is available from the corresponding author
upon reasonable request.

https://wwwn.cdc.gov/nchs/nhanes/continuousnhanes/default.aspx?BeginYear=2015

## Declarations

### Data and code availability

NHANES data are publicly available from the National Center for Health Statistics (https://wwwn.cdc.gov/nchs/nhanes). The analysis notebook, which reproduces all results, is openly available on Zenodo under an MIT license: https://doi.org/10.5281/zenodo.23267842 [18].

## Competing interests

The authors declare no competing interests. The first author is employed by CGI Technologies and Solutions; no client or proprietary data were used.

### Ethics

This study used de-identified public-use data; no additional ethics approval was required.

## References

[1] Jager KJ, Kovesdy C, Langham R, Rosenberg M, Jha V, Zoccali C. A single number for advocacy and communication—worldwide more than 850 million individuals have kidney diseases. Kidney Int. 2019;96(5):1048–1050. doi:10.1016/j.kint.2019.07.012

[2] GBD Chronic Kidney Disease Collaboration. Global, regional, and national burden of chronic kidney disease, 1990–2017: a systematic analysis for the Global Burden of Disease Study 2017. Lancet. 2020;395(10225):709–733.

[3] Centers for Disease Control and Prevention. Chronic Kidney Disease in the United States. US Department of Health and Human Services; updated March 31, 2026. https://www.cdc.gov/kidney-disease/php/data-research/index.html

[4] KDIGO CKD Work Group. KDIGO 2012 clinical practice guideline for the evaluation and management of chronic kidney disease. Kidney Int Suppl. 2013;3(1):1–150.

[5] Tangri N, Stevens LA, Griffith J, et al. A predictive model for progression of chronic kidney disease to kidney failure. JAMA. 2011;305(15):1553–1559.

[6] Tangri N, Grams ME, Levey AS, et al. Multinational assessment of accuracy of equations for predicting risk of kidney failure: a meta-analysis. JAMA. 2016;315(2):164–174. doi:10.1001/jama.2015.18202

[7] Alfego D, Ennis J, Gillespie B, et al. Chronic kidney disease testing among at-risk adults in the U.S. remains low: real-world evidence from a national laboratory database. Diabetes Care. 2021;44(9):2025–2032. doi:10.2337/dc21-0723

[8] Shin JI, Chang AR, Grams ME, et al. Albuminuria testing in hypertension and diabetes: an individual-participant data meta-analysis in a global consortium. Hypertension. 2021;78(4):1042–1052. doi:10.1161/HYPERTENSIONAHA.121.17323

[9] Xiao J, Ding R, Xu X, et al. Comparison and development of machine learning tools in the prediction of chronic kidney disease progression. J Transl Med. 2019;17:119. doi:10.1186/s12967-019-1860-0

[10] Dong Z, Wang Q, Ke Y, et al. Prediction of 3-year risk of diabetic kidney disease using machine learning based on electronic medical records. J Transl Med. 2022;20:143. doi:10.1186/s12967-022-03339-1

[11] Christodoulou E, Ma J, Collins GS, Steyerberg EW, Verbakel JY, Van Calster B. A systematic review shows no performance benefit of machine learning over logistic regression for clinical prediction models. J Clin Epidemiol. 2019;110:12–22. doi:10.1016/j.jclinepi.2019.02.004

[12] Collins GS, Moons KGM, Dhiman P, et al. TRIPOD+AI statement: updated guidance for reporting clinical prediction models that use regression or machine learning methods. BMJ. 2024;385:e078378. doi:10.1136/bmj-2023-078378

[13] Inker LA, Eneanya ND, Coresh J, et al. New creatinine- and cystatin C–based equations to estimate GFR without race. N Engl J Med. 2021;385(19):1737–1749.

[14] Pedregosa F, Varoquaux G, Gramfort A, et al. Scikit-learn: machine learning in Python. J Mach Learn Res. 2011;12:2825–2830.

[15] DeLong ER, DeLong DM, Clarke-Pearson DL. Comparing the areas under two or more correlated receiver operating characteristic curves: a nonparametric approach. Biometrics. 1988;44(3):837–845.

[16] Sun X, Xu W. Fast implementation of DeLong’s algorithm for comparing the areas under correlated receiver operating characteristic curves. IEEE Signal Process Lett. 2014;21(11):1389–1393.

[17] Van Calster B, McLernon DJ, van Smeden M, Wynants L, Steyerberg EW. Calibration: the Achilles heel of predictive analytics. BMC Med. 2019;17:230.

[18] Saxena JN, Potturu DVP, Nagraj A. Code for: Can routine clinical variables substitute for urine albumin testing in moderate chronic kidney disease? A machine-learning analysis of NHANES 2015–2018 [software], version 1.0.0. Zenodo; 2026. doi:10.5281/zenodo.23267842

[19] Inker LA, Grams ME, Levey AS, et al. Relationship of estimated GFR and albuminuria to concurrent laboratory abnormalities: an individual participant data meta-analysis in a global consortium. Am J Kidney Dis. 2019;73(2):206–217. doi:10.1053/j.ajkd.2018.08.013

